# Learning from the Experiences of COVID-19 Survivors: A Descriptive Study

**DOI:** 10.1101/2021.03.17.21253728

**Authors:** Temiloluwa Prioleau

**Affiliations:** Dartmouth College, Hanover, NH, 03755

## Abstract

**Background:** There are still many unanswered questions about the novel coronavirus, however, a largely underutilized source of knowledge are the millions of people who have recovered after contracting the virus. This includes majority of undocumented cases of the COVID-19 which are classified as mild or moderate and received little to no clinical care during the course of illness.

**Objective:** To document and glean insights from the experiences of persons with first-hand experience with coronavirus, especially the so-called mild to moderate cases that self-resolved in isolation.

**Methods:** This online-based survey study called *C19 Insider Scoop* recruited adult participants that are 18-years or older who reside in the United States and tested positive for COVID-19 or antibodies. Participants were recruited through various methods including online support groups for COVID-19 survivors, advertisement in local news outlets, and advertisement through professional and other networks. The main outcomes measured include knowledge on contraction/transmission of the virus, symptoms, and personal experiences on road to recovery.

**Results:** A total of 72 participants (53 females/19 males, ages 18 - 73 yrs. old, mean = 41-yrs.) from 22 U.S. states participated in this study. We found that the top known source of how people contracted the COVID-19 virus was through a family or household member (n=26 or 35%). This was followed by essential workers contracting the virus through the workplace (n=13 or 18%).

Participants reported up to 27 less-documented symptoms that they experienced during their illness such as brain/memory fog, palpitations, ear pain/discomfort and neurological problems. In addition, 47 out of 72 participants (65%) reported that their symptoms lasted longer than the commonly cited 2-weeks even for mild cases of COVID-19. In our study, the mean recovery time was 4.5-weeks, and exactly one-half of survivors (50%) still experienced lingering symptoms of COVID-19 after an average of 65-days following illness onset. Additionally, 37 participants (51%) reported that they experienced stigma associated with having COVID-19.

**Conclusion:** This study presents preliminary findings which suggests that emphasis on family/household spread of COVID-19 may be lacking and there is a general underestimation of the recovery time even for mild cases of the virus. Although a larger study is needed to validate these results, it is important to note that as more people experience COVID-19, insights from prior survivors can enable a more informed public, pave the way for others who may be affected, and guide further research.

## Introduction

The coronavirus (COVID-19) pandemic has significantly impacted majority of countries in the world, with a global total of more than 41.1-million confirmed cases and over 1.1 million deaths as of October 22, 2020 [1]. Months into the pandemic there are still many unknowns about the novel coronavirus, transmission, and experiences of people infected by the virus [2], [3].

However, it is known that the illness severity of COVID-19 can range from mild to critical; studies from the Center for Disease Control (CDC) in the United States and China show that around 80% of the affected population experience “mild to moderate” symptoms [4], [5]. Due to the sheer volume of affected persons and limited capacity of the healthcare systems, it was and still is recommended that patients with mild to moderate cases of COVID-19 (∼80%) manage their illness in isolation [2]. Many of these patients, especially in the early months of the outbreak (i.e., February and March 2020 in the U.S. [6]), received little to no clinical care [7]; hence there is a critical gap in knowledge about their experiences with COVID-19. According to the CDC, “recognition of factors associated with amplified spread during the early acceleration period will help inform future decisions” and “strengthen systems to detect potential transmission resurgence” [6].

Unlike hospitalized patients, the population of people with mild to moderate symptoms of COVID-19 remains largely understudied and are amongst the “undocumented masses,” yet this population contributes significantly to rapid transmission of the virus [3], [8], [9]. To fill in the gap, this work aims to enable the research and larger community glean insights from the experiences of COVID-19 survivors, especially those whose journey with the virus is not captured in clinical or medical records. Through personal stories, this study aims to bring awareness to how people in the U.S. contracted the coronavirus, the range of symptoms experienced during illness, duration of illness, and experiences on the road to recovery. Such knowledge can inform guidelines for the vast majority of cases of COVID-19 cases which are considered mild to moderate in severity, especially given the current resurgence in the United States [10]. In this work, we developed and deployed an online platform for collating experiential data from persons who recovered and/or are on the road to recovery from COVID-19. As previously mentioned, the primary aim of this work is to garner insights from persons with first-hand experience with coronavirus, especially those with mild to moderate cases which are less documented in literature. A total of 72 subjects from 22 U.S. states with laboratory-confirmed positive tests (n=68 or 94%) and presumptively positive as identified by a clinical personnel (n=4 or 6%) were recruited to share their story with COVID-19. Details on recruitment and participant characteristics are described in the methods section.

## Methods

### 1. Data Collection

This research study was approved by Committee for Protection of Human Subjects (CPHS) at Dartmouth College and is reported in accordance with the CHERRIES checklist for online surveys [11]. We recruited COVID-19 survivors defined as persons who tested positive for the coronavirus and/or who were later confirmed to have had the virus by testing positive for antibodies. Subjects were recruited through multiple sources including through online support groups such as Survivor Corps [12], advertisement in local newspapers [13], and sharing about the research study through professional and other networks. In total, 72 participants were recruited between the dates of May 10 through October 1, 2020 although majority (i.e., 54 out of 72 participants or 73% were recruited between May 10 and June 18, 2020). About half of participants in this study, 35 out of 72 (49%) were recruited from the Survivor Corps online support group following the participant’s self-identification of having COVID-19. For this population, a recruitment message was sent directly to prospective participants informing them about the study and requesting their participation if fitting. All interested participants who learned about the study were directed to the project website and invited to first complete a pre-survey on the *C19 Insider Scoop* project website [14] for screening on the eligibility requirements of being 18-years or older, living in the Unites States, and testing positive for COVID-19 and/or COVID-19 antibodies. Participants who met these eligibility requirements for the study were then provided a password by email and a link to the full-survey through which they shared their story with COVID-19. In total, 105 prospective participants completed the pre-survey and 72 participants completed the full survey thereby providing data to be presented in this study. Eligible participants who completed the study requirements (i.e., pre-survey and full-survey) were provided the option of receiving a monetary incentive for participation.

### 2. Full Survey for Sharing COVID-19 Experience

The full survey was developed using the well-established Qualtrics Survey Software [15] under Dartmouth College’s license to ensure secure data storage behind institutional firewalls. The survey questions in this study were informed by various sources including the Center of Disease Control and Prevention’s coronavirus case report form [16], the National Institute of Health (NIH) repository of COVID-19 research tools [17], and knowledge gaps identified in literature [7]. The full survey included questions covering the themes of: i) descriptive characteristics, ii) contraction/transmission, iii) symptoms and coping strategies, and iv) the road to recovery. In addition, adaptive questioning was implemented such that some questions were only displayed based on responses to previous items. On two occasions, additions were made to the full survey during data collection. More specifically, on May 21, 2020, the symptoms of headaches, muscle/body aches, and dizziness were added to the default checklist provided in this study; this was after 20 participants had completed the full survey. On May 28, 2020, questions on antibody testing were added; this was after 32 participants had completed the full survey. Persons who completed the survey before each revision have no responses to the questions added after their participation. An exact copy of full survey used in this study can be found in the appendix section of this manuscript.

### 3. Description of Study Participants

A complete demographic summary of participants is presented in Table 1. Of the 72 (53 females, 19 males) subjects, 68 (94%) received a positive lab test and/or positive antibody test for COVID-19 while 4 (6%) were confirmed presumptively positive by a clinical personnel.

**Table 1:** Demographic Summary of Participants (i.e., COVID-19 Survivors) in the C19 Insider Scoop Study (n=72)

|  |  |  |  |
| --- | --- | --- | --- |
| **Age (years)** |  | 18 – 73 (mean = 41+/- 14) |  |
| **Sex** |  | **COVID-19 Testing** |  |
|  | **<u>n (%)</u>** |  | **<u>n (%)</u>** |
| Female | 53 (74) | Positive lab/antibody test | 68 (94) |
| Male | 19 (26) | Presumptively positive | 4 (6) |
| **Race** |  | **Highest Level of Education** |  |
|  | **<u>n (%)</u>** |  | **<u>n (%)</u>** |
| White | 43 (60) | 4-year degree | 25 (35) |
| Black/African American | 14 (19) | Professional degree | 18 (25) |
| Asian | 6 (8) | Some college | 14 (19) |
| American Indian/Alaska Native | 2 (3) | High school/GED | 6 (8) |
| Other/Mixed Race | 7 (10) | Doctorate | 5 (7) |
|  |  | 2-year degree | 4 (6) |
| **U.S. State of Resident** |  | **Household Income Level** |  |
|  | **<u>n (%)</u>** |  | **<u>n (%)</u>** |
| New York | 25 (35) | \$80,000 - \$139,999 | 25 (35) |
| California | 5 (7) | \$140,000 - \$199,999 | 15 (21) |
| Georgia | 5 (7) | Less than \$20,000 | 13 (18) |
| Massachusetts | 4 (6) | \$200,000+ | 9 (12) |
| Virginia | 4 (6) | \$40,000 - \$79,999 | 9 (12) |
| Texas | 3 (4) | Not reported | 1 (1) |
| New Hampshire | 3 (4) |  |  |
| 8 other states | 2 (3) |  |  |
| 7 other states | 1 (1) |  |  |
| **Pre-existing Medical Condition** |  | **Occupation** |  |
|  | **<u>n (%)</u>** |  | **<u>n (%)</u>** |
| No, with pre-existing condition | 48 (68) | Non-essential worker | 20 (28) |
| Yes, with pre-existing condition | 23 (32) | Essential worker (Healthcare) | 14 (19) |
| Not reported | 1 (1) | Essential worker (Non-healthcare) | 14 (19) |
|  |  | Unemployed | 12 (17) |
|  |  | Student | 7 (10) |
|  |  | Retired | 5 (7) |

Participants ranged in age range from 18 to 73 years old and were residents of a total of 22 U.S. states with New York having the highest representation. The race demographics included 43 (60%) Whites, 14 (19%) Blacks/African Americans, 6 (8%) Asians, 2 (3%) American Indians/Alaska Natives and 7 (10%) other/mixed race. In addition, a total of 23 participants (32%) had at least one pre-existing medical condition and 6 (8%) had more than one pre-existing medical condition. The most common pre-existing conditions reported include asthma (n=15), high blood pressure (n=4) and immunosuppressive conditions (n=3). Other descriptive factors such as highest level of education, household income and occupation are detailed in Table 1.

## Results

### 1. Onset, Testing, Contraction of COVID-19

Figure 2 (A) shows that 51 participants in this study (i.e. 72%) experienced COVID-19 symptom-onset in March of 2020, about 40 days after the first reported case in the U.S. [18]. Out of the total of 72 subjects, 30 (42%) were working from home at the time of their symptom onset while 42 (58%) were not working from home. In addition, 46 (64%) reported to have been using recommended precautionary measures prior to the onset of their symptoms such as masks (n=25), social distancing (n=22), frequent handwashing (n=14), and gloves (n=12). However, it is important to note that majority of participants experienced symptom-onset during the first accelerated spread of COVID-19 in the U.S., in March 2020, at which time recommendations for personal protective practices was only beginning to unfold [6]. In retrospect, 63 participants (87%) could narrow down the source of how they might have contracted the COVID-19 virus; 62% shared probable sources while 25% shared what they consider to be definite sources of their infection. From the reported data shown in Fig. 2 (B & C), the most prevalent source of virus contraction was from a family or household member. More specifically, 26 participants (36%) attributed the source of their virus to a family or household member. Additionally, 27 participants (38%) reported that at least one other person in their family or household contracted COVID-19 from them. This suggests that there is a moderate probability of the coronavirus spreading within persons sharing a living space even when the appropriate precautions are taken to minimize the risk. However, emphasis on family/household transmission of COVID-19 is limited. Following this, the second most prevalent source of virus transmission was essential worker who contracted the virus while on the job. This was reported by 13 participants (18%).

**Fig. 1.**
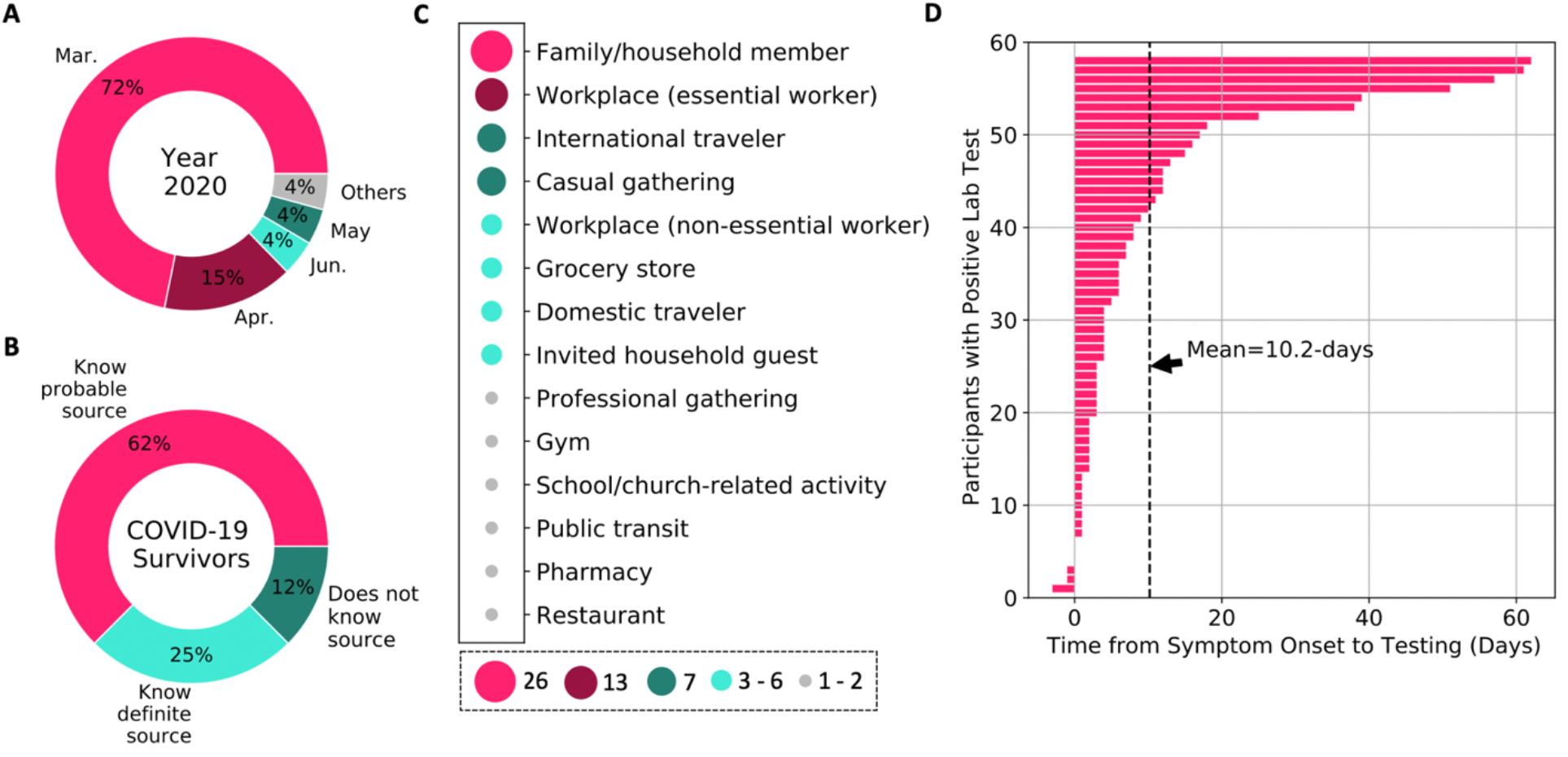
(**A**) Month of symptom-onset; (**B**) knowledge about source of infection; (**C**) prevalent sources of virus transmission, and (**D**) time lag from symptom-onset to testing.

**Fig. 2.**
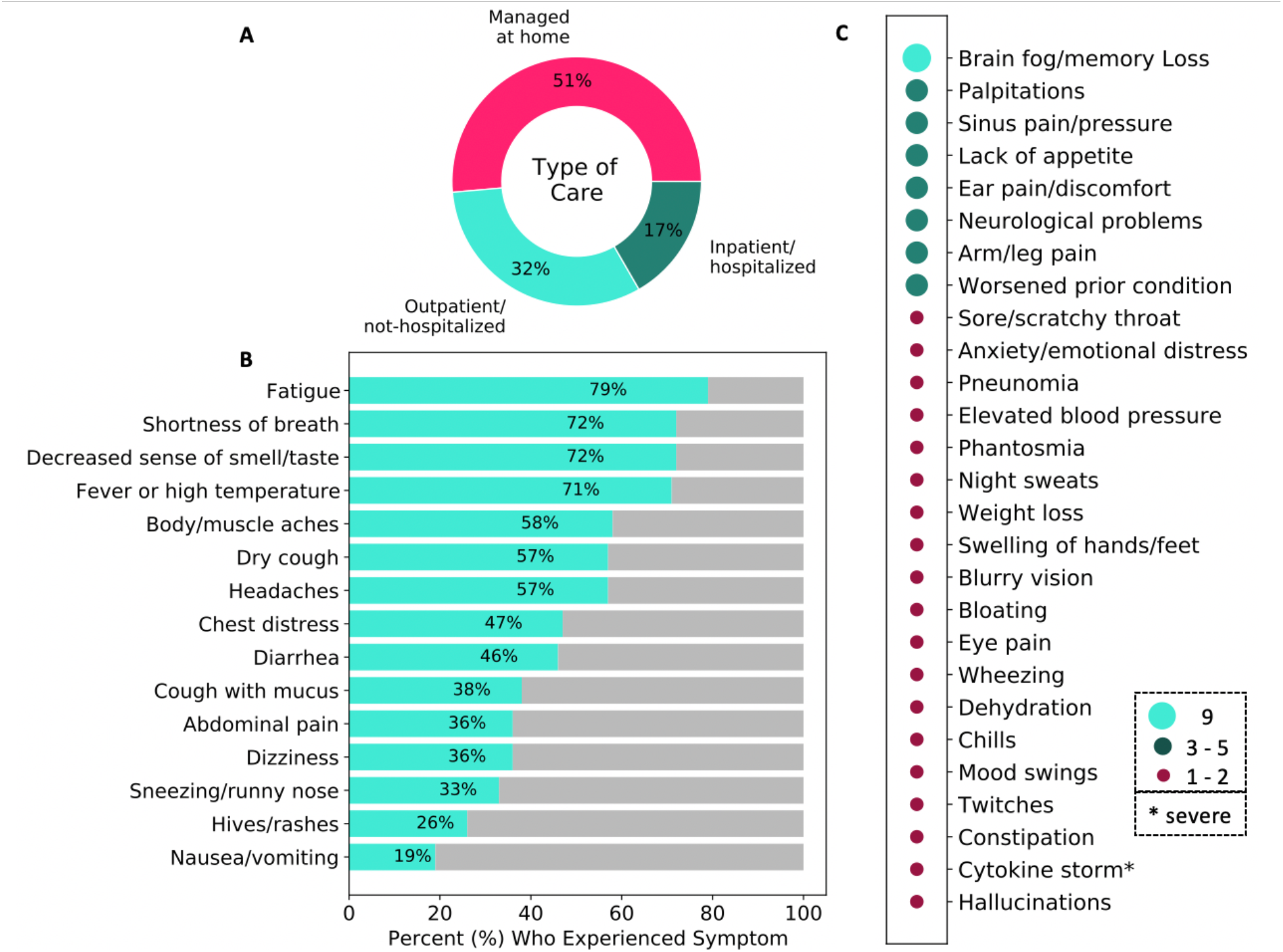
(**A**) The type of care received by COVID-19 survivors; (**B**) list and prevalence of more common symptoms; (**C**) other less-documented symptoms of coronavirus.

These two sources of virus transmission were also the top sources identified from persons who shared more definite sources of their infection. Results obtained shows other sources of virus transmission such as casual gatherings (n=7 or 10%), workplace for non-essential workers (n=6 or 8%), and even the grocery store (n=4 or 6%).

During the early months of the outbreak in the U.S., limitations in testing is identified as one of the multiple factors that contributed to the rapid spread [6], [8]. Fig. 1 (D) shows that following infection, participants experienced large variability in the time it took to gain access to testing; this ranged from 3-days before symptom-onset to 62-days after symptom-onset in this study. The average time from symptom-onset to testing was 10.2-days with about 10% of participants getting tested more than 35-days after their symptoms began. Although this research required a COVID-19 positive test for participation, we learned during recruitment that a large number of people were continuously denied access to testing during their illness. For example, a patient-led study with over 600 participants who experienced COVID-19 symptoms show that “47.8% were either denied testing or not tested for another reason” [19]. The delay in testing and uncertainty that persons with COVID-19 symptoms experienced is a possible contributor to further spread of the virus. In addition, a few participants reported that were treated for sinus infections, bronchitis, and other conditions without testing, as a result they may not have isolated early in their journey with the virus due to lack of knowledge.

### 2. A Deeper Look at COVID-19 Symptoms

There is no shortage on the characteristics of COVID-19 for severe cases in hospitalized patients [5], [7], [20], [21]. However, severe cases account for only about 20% of the full population of confirmed coronavirus cases and differences have been identified between mild and severe cases [2], [22]. Therefore, it is critical to also understand the journey of non-hospitalized persons with COVID-19 who account for 80% or more of confirmed cases.

Majority of participants in this study, 60 out of 72 (83%) managed their symptoms at home and/or in isolation – see Fig. 3 (A) ; these participants represent people with mild to moderate cases of COVID-19. This is alignment with the estimates in literature that about 80% of the COVID-19 cases are mild to moderate as opposed to severe [2], [21]. Participants were asked to identify all symptoms they experienced during their illness with COVID-19. Fig. 3 (B) shows the prevalence of recognized COVID-19 symptoms amongst this study population (n=72) with the top 4 being fatigue (n=57 or 79%), shortness of breath (n=52 or 72%), decreased sense of smell/taste (n=52 or 72%) and fever (n=51 or 71%). These more common symptoms have also been identified in earlier studies, albeit with different percentages for frequency of occurrence in various populations [21], [23], [24]. More interestingly, in addition to the well-recognized symptoms of COVID-19, 30 participants (41%) shared other less-documented symptoms that they experienced during their illness. Fig. 3 (C) shows a list of other patient-reported symptoms associated with COVID-19 and the prevalence of each amongst the population. The most prevalent from these patient-reported symptoms include brain fog/memory loss which was reported by n=9 (13%), palpitations (i.e., elevated heart rate) and sinus pain/pressure, both reported by n=5 (7%), followed by lack of appetite, ear pain/discomfort, and neurological problems, reported by n=4 (6%). Many of these other symptoms have not been emphasized in prior work. From the list of 27 additional patient-reported symptoms, cytokine storm is considered the most severe and can lead to rapid deterioration, multiple organ failure, and mortality [25]–[27]. Cytokine storm is characterized by a lethal overreaction of the immune system in response to an infection or disease. The single participant who experienced cytokine storm (Male, 62yrs) was tested for COVID-19 on April 4, 2020 and then examined by a registered nurse, however, signs of the ensuing havoc were not identified. Following clinical evaluation, the participant was guided to self-quarantine, however within 72-hours he was taken to the hospital as “COVID-19 had triggered a massive cytokine storm.” He was hospitalized for 36-days, on the ventilator for 9-days, and needed surgery to repair the collapse of his left lung due to COVID-19. Fortunately, he lives to tell his story and strongly urges that COVID-19 patients should be evaluated, early in their illness and possibly during testing, for signs of cytokine storm to prevent rapid deterioration and save lives.

**Fig. 3.**
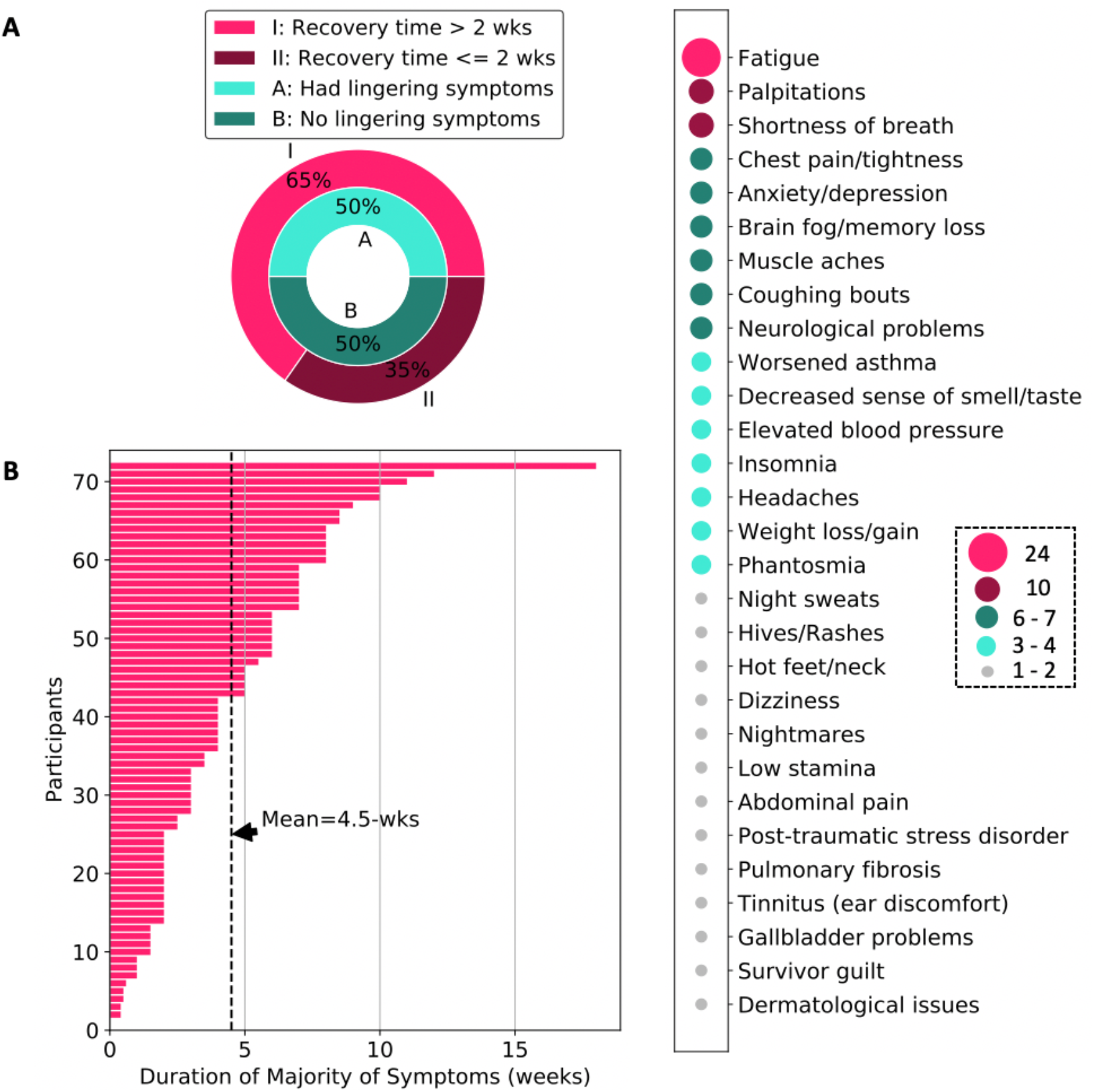
(**A**) The percent of COVID-19 survivors who experienced lingering symptoms and longer than the commonly cited 2-weeks recovery time; (**B**) duration of majority of symptoms across study participants showing a mean of 4.5-weeks; (**C**) list and prevalence of lingering symptoms associated with coronavirus.

### 3. The Road to Recovery from COVID-19

There are many unknowns about the road to recovery for persons who contract COVID-19. According to a February 2020 remark from the World Health Organization (WHO) director-general, the recovery time for persons with mild cases of coronavirus is about 2-weeks and 3 to 6-weeks for persons with severe or critical cases [28]. However, results from our study show that 47 out of 72 participants (i.e., 65%) experienced longer than a 2-week recovery time – see Fig. 4 (A). More specifically, we found that the mean time to recover from “majority of symptoms” across all participants was 4.5-weeks; the average recovery time for non-hospitalized patients (i.e., persons with mild to moderate severity) was 4.2-weeks while the recovery time for hospitalized patients (i.e., persons with more severe experience of COVID-19) was 6.2-weeks or more. A few hospitalized patients reported that they had not recovered from majority of their symptoms at the time of participating, thus the length of their illness at time of participation was used as a proxy (i.e., date of participation minus date of symptom onset). Our study also shows that COVID-19 survivors experienced high variability in their recovery time from the illness, ranging from 0 days - reported by asymptomatic patients to 18 weeks - reported by a 58-year-old female with one pre-existing condition and three hospital visits for COVID-19 related symptoms including dehydration and vestibular migraine.

**Fig 4.**
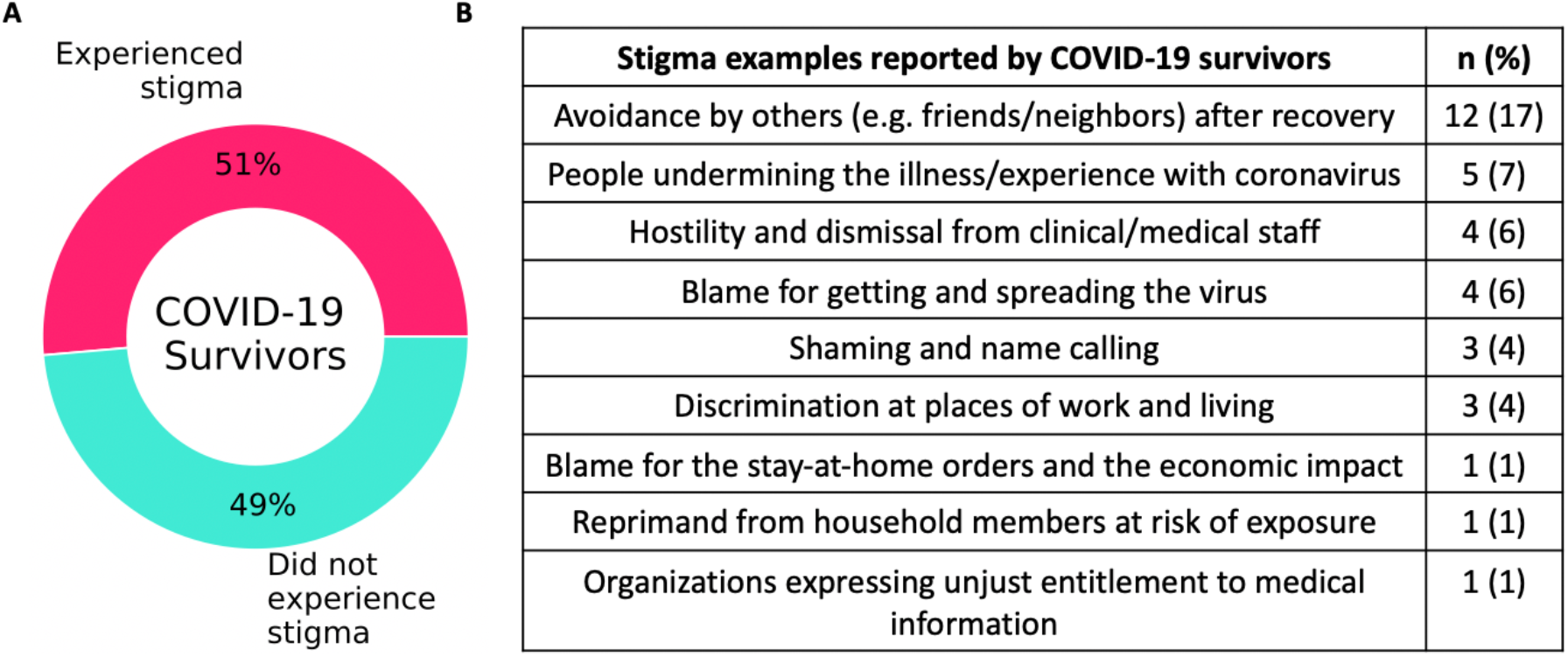
(**A**) The percent of COVID-19 survivors who experienced stigma following their illness; (**B**) examples of prevalent stigmas reported.

In addition to the longer recovery times identified in this work, we found that 36 out of 72 participants (50%) still experienced lingering symptoms of COVID-19 even after an average of 65-days following their illness onset. Fig. 4 (C) shows a list lingering symptoms and changes to general health identified by participants from this study. The most common issues identified include fatigue reported by 24 participants (33%), followed by shortness of breath and palpitations reported by 10 participants (14%), then chest pain/tightness, anxiety/depression, brain fog/memory loss, muscle aches, and coughing bouts, all reported by 6 – 7 participants (8 – 10%). Out of the 72 participants, 37 people (51%) reported to have experienced stigma following their contraction of COVID-19 while 35 people (49%) reported otherwise. As shown in Fig. 5, there were 9 themes that stood out from further descriptions of the stigmas experienced by persons who contracted COVID-19. The most prevalent stigma was avoidance by others after recovery – reported by 12 participants (17%) followed by people undermining the illness/experience with coronavirus – reported by 5 participants (7%). Other examples of stigma reported include hostility and dismissal from clinical/medical staff and being blamed for getting and spreading the virus.

### 4. Insights from COVID-19 Survivors

COVID-19 survivors are an invaluable resource not being fully utilized as a source of knowledge. Through this study, survivors shared insights that they believe will help the society at large better cope with the realities of ongoing pandemic. Many of these insights are targeted toward the larger population who may not yet have been infected by the coronavirus, however, with the recent rise in COVID-19 cases in the U.S. and other countries [10], these insights are critical for the general public.

Salient insights from COVID-19 survivors are as follows (direct quotes from participants are presented in quotation marks):

1. “Any symptom can be the virus so self-isolate even if you have a tickle in your throat… It is easy to mistake a mild case of COVID-19 for a cold or allergies” in the early stages.
2. Understand the “fact that doctors and nurses do not have all the answers” so set your expectations accordingly.
3. The journey with COVID-19 can be scary and lonely as well as physically, mentally, and emotionally tasking. “You have to advocate for yourself every step of the way.”
4. Young and healthy people without any underlying medical conditions can also have a rough journey with the virus. Take it seriously. Protect yourselves and others.
5. “The disease presents differently in all patients. It is not a *one size fits all*.” For some it is short-lived while others suffer long-term health issues as a result of the COVID-19.
6. If you contract the virus, take it day by day. Be patient with the recovery process as it can be long and volatile. Symptoms and lingering symptoms can come in waves.
7. Do not wait too long to go to the hospital (if needed) as a delay has led to worse outcomes for some. Additionally, some patients have gone to the hospital and been dismissed or discharged pre-maturely. Trust your instinct as the sole person with first-hand knowledge of your own illness.
8. Do not allow distrust of the health care system to be your excuse for having a worse outcome. Ask questions during clinical care to ensure that you are being prescribed the best treatment for your specific situation.
9. Some medications used in severe cases can cause hallucination and uncanny dreams.
10. Stigma during or after COVID-19 is a reality. Others may have various reactions to people who contracted the virus, especially during the initial phase of returning back to society. Some people may be cruel and unkind with their actions or words.
11. COVID-19 tests and antibody tests can be inaccurate. There are persons who test positive for COVID-19 and negative for antibodies, and vice versa.
12. It is not sufficient for organizations to rely on body temperature as a primary means for determining COVID-19 symptoms. Some survivors never experienced fever while others only experienced fever for a short time (e.g., 24-hours).

## Discussion

This paper presents a descriptive study that summarizes the experiences of first-hand COVID-19 survivors with a particular focus on how people contracted the virus, the range of symptoms, the duration of illness, and experiences on the road to recovery. Our findings show that the top known source of how people contracted COVID-19 was through a family or household member. Participants also reported a range of symptoms include up to 27 less-documented symptoms that they experienced during their illness such as brain/memory fog, palpitations, ear pain/discomfort and neurological problems. Another key finding is that majority of participants (47 out of 72 or 65%) experienced longer than the commonly cited 2-weeks recovery time. The mean recovery time in this study was 4.5-weeks; more specifically 4.2-weeks for non-hospitalized patients (i.e., mild to moderate cases) and 6.2-weeks or more for severe cases (i.e., patients that were hospitalized for COVID-19). In addition, a little over half of participants (n=37 or 51%) reported that they experienced stigma associated with having COVID-19. Examples of stigmas shared include avoidance by others after recovery and people undermining their illness/experience with the coronavirus. Finally, participants shared insights from their personal journey with COVID-19 in hopes to inform and encourage the general public, especially others who may not yet have been infected by the coronavirus.

### 1. Comparison with Prior Work

Majority of early literature that presented characteristics of COVID-19 focused on persons with severe cases of the virus, primarily hospitalized patients [5], [21], [23], [24]. This created a critical gap in literature on the majority of COVID-19 cases (∼80%) which are categorized a mild or moderate (i.e., non-hospitalized cases). One of the first questions relating to the COVID-19 pandemic is “how is it spreading amongst people?” Early sources of virus transmission in the U.S. was primarily from international travelers returning to the country such as the “First [Reported] Case of 2019 Novel Coronavirus in the United States” [18]. However, community spread became prevalent not too long after. Later reports by the Center for Disease Control and Prevention (CDC) identified transmission through a family member or work colleague as a prevalent source of virus contraction as well as transmission through social gatherings such as dining at a restaurant [29], [30]. Results from this study show agreement that family/household- and work-related transmission is prominent.

Other studies have now begun to uncover characteristics of mild to moderate cases of COVID-19. For example, the work by Liu et al. reported viral shedding patterns observed in patients with mild and severe COVID-19 [22]. The authors identified some key differences between patients with mild and severe cases of COVID-19 thereby supporting the need for studying both populations comprehensively. Xu et al. [31] presented a telemedicine system to continuously monitor the progression of home-quarantined patients with COVID-19, 74 of whom had confirmed cases of the virus. They identified symptoms and trajectories that were indicative of need for hospitalization. More closely related to this work, Jeon et al. [32] used a combination of biomedical literature and social media data to characterize symptoms of COVID-19. The authors identified 25 novel symptoms through social media posts, specifically Twitter posts. Some of the less common symptoms identified in that work align with the less-documented symptoms reported by participants in this study such as ear and eye problems, weight loss, and memory disorder, however, results in this study also include symptoms that were not identified by Jeon et al. [32] such as worsened prior condition (e.g., asthma) and elevated blood pressure.

Even less studies in literature have focused on understanding the time to recovery after illness with COVID-19. One of the earliest sources of information on recovery time was a February 2020 remark from the WHO director which stated that the recovery time for persons with mild cases of coronavirus is about 2-weeks and 3 to 6-weeks for persons with severe or critical cases [28]. Subsequently, a study from CDC found that approximately 35% of symptomatic outpatients with COVID-19 had not returned to baseline health 14 – 21 days after test date [9]. Carfi et al. assessed persistent symptoms in patients after acute COVID-19 and found that 87.4% of patients with severe cases of COVID-19 reported at least 1 symptom, particularly fatigue and dyspnea, after an average of 60-days from symptom onset [33]. Results from this study found the average recovery time for mild cases is 4.2-weeks and 6.2 weeks or more for severe cases. This is in alignment with the findings from the CDC report [9] and Carfi et. al [33], however, our results provide concrete numbers based on participants in this study.

### 2. Limitations

There are several limitations in this work. Firstly, the sample size of the study is small compared to the total number of people in the U.S. that have contracted COVID-19. However, this study benefits from diverse recruiting strategies and thus has representation from 22 out of the 50 states in the country. A second limitation is that this study relies on participant’s ability to recall and share their experience with COVID-19. There are well-known limitations of self-report-based studies such as recall bias. However, we believe this is minimized because majority of subjects in this study experienced symptom onset in March 2020 and were recruited between the months of May and June 2020, this means that participants were recruited about 2-months after they contracted the virus. In addition, the average duration of illness was 4.5-weeks across all subjects and participation in this study required recovery from the illness, hence we expect that majority of subjects participated within their first month after recovery thus maximizing their chance to accurately remember their experience.

### 3. Conclusion

It is well-known that many people do recover from COVID-19, however, results from this study show that the journey for some may be long and uncomfortable. To support an accurate depiction of the journey with COVID-19, it is important to recognize that the recovery time can be more than 2-weeks for mild symptoms. In addition, a notable percent of COVID-19 survivors are considered “long haulers” [34], [35]. This refers to people who experience lingering symptoms weeks and even months after initial contraction. More research is needed to understand the reasons for such diverse experiences with the novel coronavirus. Also, thousands of people have died from the same virus with disproportionate percentages amongst Black and Hispanic populations in the United States [36]. More research is needed to better understand the trajectory of cases that ended in fatality and help prevent a similar outcome for others. One of such efforts should be toward early detection of cytokine storm triggered by the coronavirus as this has been shown to cause fatality for some of the affected population.

## Supporting information

Survey Questions

## Data Availability

The data used in this study is not made publicly available to protect the privacy of participants.

## Acknowledgements

The author acknowledges the many participants and advocates of the *C19 Insider Scoop project* listed on www.c19insider.com with special thanks to S. Prioleau for designing and building the project website for data collection. This work is supported by Dartmouth COVID-19 ‘Spark’ funding and a “RAPID” grant from the National Science Foundation (NSF) – award number: 2031546. The author T. Prioleau conceived the study, developed the survey, curated the data, performed analysis, and wrote the manuscript. This author has full access to all the data in this study and takes responsibility for the integrity of the data and accuracy of the data analysis. T. Prioleau has no competing interest to declare.

## Notes

### Competing Interest Statement

The authors have declared no competing interest.

### Author Declarations

This research study was approved by Committee for Protection of Human Subjects (CPHS) at Dartmouth College.

