## Supplementary material for "Learning from the Experiences of COVID-19 Survivors: A Descriptive Study": Survey Questions

### Welcome

Welcome and thank you for choosing to share your voice and experience with COVID-19. This is one way you can contribute in the fight against this global pandemic. Note that the survey data is anonymous and your information will be maintained with the highest priority. The estimated completion time is 30 mins. Feel free to check the project website (c19insider.com) periodically for the insights we are learning.

### Create an alias

Please create an alias name that will be used as your participant ID and for acknowledgement (e.g. MP32 for Mary Poppins). If you filled out the pre-survey, use the same alias so that your incentive can be processed.

If relevant, please tell us who referred you by using the first and last initial for specific people (e.g. KD). If referred by or associated with an organization, simply type the name (e.g. 'survivor corps', 'valley news', etc.).

### About You

### Gender

- ○ Male
- ○ Female
- ○ Other

### Year of Birth (e.g. 1978)

### Ethnicity

- ○ Hispanic or Latino
- ○ Not Hispanic or Latino

### Race (check all that apply)

- ☐ Asian
- ☐ Black or African American
- ☐ White
- ☐ American Indian/Alaska Native
- ☐ Native Hawaiian/Other Pacific Islander
- ☐ Unknown
- ☐ Other

### Do you live in the United States?

- ○ Yes
- ○ No

State of residence

County of residence (e.g. "Fulton" county in Atlanta, "Harris" county in Texas)

Highest-level of education completed

Current living situation

Please describe your current living situation

Number of people in your household

Household income level

Occupation/Employment

[ ]

What is your specific role at work? (e.g. pediatric nurse, grocery attendant, high school teacher)

[ ]

### Health questions

Do you currently have health insurance?

- ○ Yes, I have health insurance
- ○ No, I do not have health insurance

Who pays for your health insurance? Check all that apply.

- ☐ Self-funded
- ☐ Current employer
- ☐ Former employer
- ☐ Government program
- ☐ Other

Do you have any pre-existing health conditions? (e.g. asthma, diabetes, hypertension)

- ○ Yes
- ○ No

### What pre-existing health condition(s) do you have? Check all that apply

- ☐ Asthma
- ☐ Obesity
- ☐ Type 1 diabetes
- ☐ Type 2 diabetes
- ☐ High blood pressure
- ☐ Cancer in the past year
- ☐ Chronic kidney disease
- ☐ Chronic lung disease
- ☐ Chronic heart disease
- ☐ Immunosuppressive condition
- ☐ Other

### List any other pre-existing health conditions that were not mentioned in the previous question

### Have you seen a healthcare provider in the last 24-months?

- ○ No, I have not
- ○ Yes, at least once
- ○ Yes, multiple times

### Have you been to the emergency room in the last 24-months?

- ○ No, I have not
- ○ Yes, at least once
- ○ Yes, multiple times

What was the reason for your visit to the emergency room?

Have you had the flu shot in the last 24-months?

- Yes, I got the flu shot earlier this year or last year
- Yes, I got the flu shot two years ago
- No, I did not get the flu shot in the last 24-months

### Testing

Did you test positive for COVID-19?

- Yes, I had a lab test that came back positive
- No, but I suspect I had COVID
- No, I did not have COVID at any point

Why do you believe that you had COVID-19?

When did you start to feel ill or experience symptoms? MM-DD-YY

When did you take the COVID test? If you don't remember use an approximate date. MM-DD-YY

How long did it take to receive the results from your test for COVID-19?

- Less than 24-hours
- 24-hours to 3-days
- Greater than 3-days

Were you working from home before you showed symptoms and/or tested positive for COVID-19?

- Yes
- No

When did you start working from home? If you don't remember use an approximate date. MM-DD-YY

Were you using any precautionary measures before you contracted COVID-19? If so, please share what measures you had in place (e.g. strict adherence to social distancing, frequent use of masks in public spaces etc.)

Did you go out of your home between when started to feel ill and when you received the positive test result?

- Yes, to take care of obligations (e.g. work, grocery shopping)
- Yes, but only for a brief time (e.g. taking a walk)
- No, I strictly self-isolated in my home
- Other

What places do you remember visiting between the time you started for feel ill and when you received the positive test result? (e.g. work, grocery store, hair salon)

What is your primary means of transportation?

- Personal vehicle
- Public transportation (e.g. train or bus)
- On foot

### Contraction/Transmission

Do you know how you contracted COVID-19?

- Yes, I definitely know when/how

- ◯ I am not sure but I have a suspicion
- ◯ No, I have no clue

Please share as much detail as possible regarding how you contracted COVID-19. Include location, event title, dates, number of persons you came in contact with, etc. Use words like, I am sure..., I suspect..., I do not know but here is some additional details...

Did you go out of your home at any point after you received a positive test and before you fully recovered?

- ◯ Yes, to take care of obligations (e.g. work, caring for others)
- ◯ Yes, but only briefly and nearby (e.g. taking a walk)
- ◯ No, I strictly self-isolated in my home

Do you know of any others who may have contracted COVID-19 from you?

- ☐ Yes, others in my household or family
- ☐ Yes, others in my workplace or school
- ☐ Yes, others in my neighborhood or vicinity
- ☐ No, I do not believe anyone else got sick from me

What is your estimate of the number of people who may have contracted COVID-19 from you? Note: This question is strictly to inform research on the infection rate of COVID-19. These are anonymous answers and the question can be skipped entirely.

[dropdown]

### During illness

Did you experience any symptoms during the course of your illness?

- ○ Yes, severe symptoms
- ○ Yes, mild symptoms
- ○ No symptoms

What symptoms did you experience? Check all that apply.

- ☐ Fever or high temperature
- ☐ Dry cough
- ☐ Cough with mucus
- ☐ Sneezing/runny nose
- ☐ Fatigue
- ☐ Shortness of breath
- ☐ Abdominal pain
- ☐ Decreased sense of smell/taste
- ☐ Chest distress
- ☐ Diarrhea
- ☐ Nausea/vomiting
- ☐ Hives/rashes
- ☐ Headaches
- ☐ Body/muscle aches
- ☐ Dizziness

☐ Others

What other symptoms did you experience?

Did you visit a hospital or clinical center for treatment during the course of your illness?

○ Yes, I was hospitalized
○ Yes, but I was not hospitalized
○ No, I managed myself at home

How many days were you hospitalized for? (e.g. 10-days)

Were you put on a ventilator while hospitalized?

○ Yes
○ No

How many days were you on the ventilator? (e.g. 7-days)

What home treatments or remedies did you use during your illness? List as many things as you believe aided your recovery.

What were your coping strategies during your illness? List anything that you believe helped you cope during your illness.

### Recovery

Are you fully recovered from COVID-19?

- Yes, I am fully recovered with no more symptoms
- Mostly, but I still have a few lingering symptoms
- No, I still have several symptoms

Please list the lingering symptoms that you currently have?

When do you believe you fully recovered from COVID-19 and associated symptoms? MM-DD-YY

How many weeks did it take you to recover from majority of the symptoms associated with COVID-19?

Have you taken a COVID-19 antibodies test?

- Yes
- No

When did you take the COVID-19 antibodies test? MM-DD-YY

What was the result of the antibodies test?

- Positive for antibodies
- Negative for antibodies

Have you experienced any stigma associated with having COVID-19?

- Yes
- No

Please share more about your experience with stigmas associated with having COVID-19?

Are there any changes to your general health after recovering from COVID-19? If so, please share.

### Mortality

Do you personally know anyone who died from COVID-19 and/or related complications?

- Yes, I know at least 1 person
- No, I do not know anyone

What is your relationship with the above person(s) who died from COVID-19 and/or related complications?

- Immediate family member (e.g. mother, father, brother or sister)
- Extended family member (e.g. cousin, aunty, uncle)

- ○ Friend
- ○ Co-worker
- ○ Other

Did the above person(s) have any pre-existing conditions?

- ○ Yes, 1 or more pre-existing conditions
- ○ No, they had no pre-existing conditions
- ○ I do not know if they had any pre-existing conditions

What pre-existing conditions did the above person(s) have? Name all that you're aware of and leave blank if you don't know.

It is known that COVID-19 deaths are disproportionately higher in certain populations. With that in mind, do you have any insights or perspective as to why the above person(s) who died from COVID-19 may have had a fatal outcome? Please share as much details as possible about the specific person(s) story (including their gender, age, race and journey with COVID-19).

**Perspective questions (Likert-Scale)**

Please state how much you agree or disagree with statements in this category.

I trust that my healthcare provider has my best interest in mind and gives me the best medical care and information available.

- Strongly agree
- Agree
- Neutral
- Disagree
- Strongly disagree

I believe the recommendation to abide by social distancing and wear a mask is appropriate and helpful to minimize the risk of contracting COVID-19.

- Strongly agree
- Agree
- Neutral
- Disagree
- Strongly disagree

Given my personal circumstance (e.g. with work, living, and health), it is easy for me to follow the recommendation of social distancing and wearing a mask to minimize the risk of spreading/contracting COVID-19.

- Strongly agree
- Agree
- Neutral
- Disagree
- Strongly disagree

### Farewell

Are there any other comments or insights that you want to share with regards to your experience with COVID-19?

We thank you again for your time and insight. Sincerely your researchers at Dartmouth College!

Powered by Qualtrics
